# The effect of secretor status and the vaginal microbiome on birth outcome

**DOI:** 10.1101/2021.11.24.21266804

**Authors:** S Kundu, YS Lee, L Sykes, D Chan, H Lewis, RG Brown, L Kindinger, A Dell, T Feizi, S Haslam, Y Liu, JR Marchesi, DA MacIntyre, PR Bennett

## Abstract

Mutations in the *FUT2* gene that result in a lack of expression of histo-blood group antigens on secreted glycoproteins may shape the vaginal microbiota with consequences for birth outcome. To test this, we analysed the relationship between secretor status, vaginal microbiota and gestational length in an ethnically diverse cohort of 313 pregnant women, including 91 who delivered prematurely. *Lactobacillus* species were found to co-occur less often with other microbial taxa in non-secretors. Moreover, non-secretors with *Lactobacillus* spp. depleted vaginal microbiota in early pregnancy had significantly shorter gestational length than *Lactobacillus* spp. dominated non-secretors (mean of 245.5 (SD=44.5) versus 265.9 (23.6)); p=0.045), but not compared to *Lactobacillus* spp. dominated (261.8 (27.5)) and depleted (264.3 days (21.2)) secretors. In identifying a relationship between blood-group antigen expression and vaginal microbiota-host interactions, our results point towards stratification by secretor status as an important factor for considering preterm birth risk and prevention.

## Introduction

Histo-blood group antigens (HBGA), ABH and Lewis, are carbohydrate sequences found on the surface of a range of cell types and in secretions such as cervico-vaginal fluid. These antigens can serve as attachment sites and energy sources for microbial organisms ^1–3^. The *FUT2* gene encodes the α(1,2)-fucosyltransferase enzyme, which adds a fucose residue to the terminal galactose on a type 1 glycan precursor forming the H antigen. Four mutations in the *FUT2* gene have been identified (alleles *se*^*302*^, *se*^*385*^, *se*^*428*^ and *se*^*571*^) that result in non-secretion of these antigens from mucosal surfaces, in homozygotes, and up to 20% of human populations express this non-secretor phenotype ^4–7^. Non-secretors will present as either Le(a+b-) or Le(a-b-), depending upon the presence of mutations in their *FUT3* gene. Secretor status has been associated with increased and decreased susceptibility to a range of bacterial and viral pathogens as well as gut microbiota composition ^8–14^.

The vaginal microbiome plays an important role in influencing pregnancy outcome. Dominance of the vaginal niche by *Lactobacillus* species has been widely reported to associate with healthy, term pregnancy ^15,16^. In contrast, high diversity microbial communities depleted of *Lactobacillus* species and enriched with pathobionts and/or bacterial vaginosis (BV)-associated bacteria, increase the risk of adverse pregnancy outcomes including miscarriage and preterm birth ^17–19^. Preterm birth represents a significant global health burden – it is the leading cause of death in children under age five and associates with serious short and long-term morbidities in survivors ^20–22^. Significant risk factors for spontaneous preterm birth in our populations include history of late miscarriage or preterm birth, and previous cervical excisional treatment, an operation to treat cervical pre-cancer ^23,24^. Both of these risk factors have been linked with characteristic compositions of the vaginal microbiota ^15,16,25,26^.

In a recent study of 300 pregnant women (of whom 28 experienced preterm birth), maternal secretor status was reported as an independent risk factor for preterm delivery (Caldwell et al., 2020). Specific human milk oligosaccharides (HMOs) in blood and urine have also been recently associated with vaginal microbiota in a small cohort (n=60) of women presenting with threatened preterm labour (Pausen et al., 2020). We postulated that the composition of the vaginal microbiota in pregnancy might be influenced by maternal secretor status, and that this may influence pregnancy duration. To characterise the effect of secretor status on the vaginal microbiome and gestational length, we sequenced the second exon of the *FUT2* gene to infer secretor status and undertook metataxonomic analysis of vaginal samples collected longitudinally from a cohort of 313 pregnant women, of which 91 delivered prematurely.

## Results/Discussion

### Distribution of non-secretors in our cohort

A total of 313 women were included in our study (Supplementary Table S1), of which 91 experienced spontaneous preterm birth before 37 weeks (259 days) of gestation. The ethnic distribution of the cohort was similar to the expected background prevalence within the clinical population. In total, 87 women were identified as non-secretors and 141 as heterozygous at these loci. Although nine women were heterozygous at multiple non-secretor loci, we did not identify any who were multiply homozygous. All four known non-secretor mutations were identified in our cohort. The *se*^*428*^ nonsense mutation accounted for 89.7% of the heterozygotes and 94% of the non-secretor homozygotes and was present across all ethnicities except East Asian patients. The remaining mutations had more restricted distributions: five women of East Asian ethnicity were homozygous for the *se*^*385*^ non-secretor allele (and a further five who were heterozygous) and only one *se*^*302*^ homozygote was identified in a woman of Central-South Asian descent (ten other women, of similar ethnicity, were heterozygous). We did not identify any homozygotes of *se*^*571*^. These patterns are consistent with previously documented distributions of non-secretor genetic diversity ^5–7^. Overall, 28% of the cohort were identified as non-secretor with the highest proportion in Afro-Caribbean women, 34.6%, compared with 26.8% and 19.2% in European and Central-South Asian ethnicities respectively. For all subsequent analyses secretor homozygotes were combined with heterozygotes into a single group since they are phenotypically similar ^27^.

### Lactobacilli are more refractory to co-colonisation in non-secretors

Vaginal microbiota were classified into one of five Community State Types (CST) using the ‘predominant taxon’ rule, which is concordant with clustering based methods and allows comparisons across datasets ^28,29^. The most prevalent CST observed in our cohort was CST 1 (majority *L. crispatus*) in 39.7, 45.9 and 41.1% of the patients at early, mid and late gestation respectively, followed by CST 3 (majority *L. iners*) in 30.0, 23.1 and 27.7%, CST 4 (diverse) in 14.3, 12.8 and 16.6%, CST 2 (majority *L. gasseri*) in 10.1, 10.7 and 7.5% and CST 5 (majority *L. jensenii*) in 5.9, 7.5 and 7.1%.

In samples taken during early pregnancy, prior to any interventions, we observed a similar prevalence of major bacterial genera across secretor and non-secretor women, including those classically associated with BV (a clinical syndrome of vaginal discharge and odour characterised by polymicrobial overgrowth), e.g., *Gardnerella, Atopobium, Anaerococcus* and *Sneathia*, (Figure 1A). As expected, most women had a high relative abundance of *Lactobacillus*, though in the secretors there was evidence of a greater spread (Figure 1B). At species level, an enrichment for CST 3 in non-secretors and CST 4 in secretors was observed but these differences were not statistically significant (Fisher’s Exact test p=0.163, Figure 1C).

**Figure 1:**
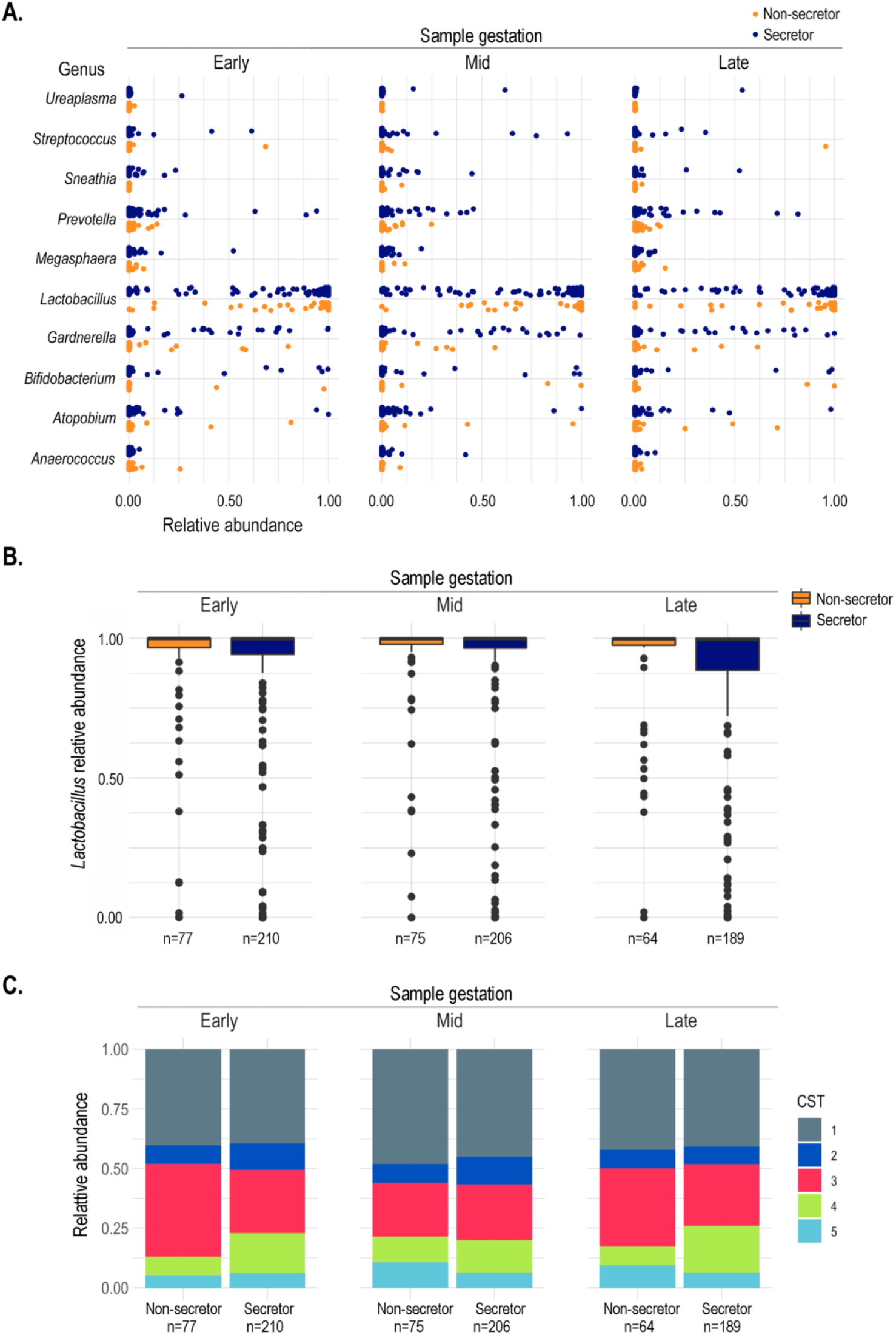
A. Distribution of the top 10 most abundant bacterial genera in the vaginal microbiota of secretors and non-secretors in early, mid and late pregnancy. B. The distribution of the proportion of *Lactobacillus* in secretors and non-secretors in early, mid and late pregnancy was similar. C. Frequencies of each Community State Type (CST) through pregnancy in secretors and non-secretors.

We observed no difference in bacterial diversity (Shannon Index) between non-secretors and secretors in early pregnancy (p-value=0.409) nor any differential classification in principal components analysis (PCA) (p-value=0.303). Differential abundance analyses (DAA) suggested no significant differences of relative abundance in species on the basis of secretor phenotype (Supplementary Figure S1) however, the low prevalence of *Lactobacillus* depleted patients (26.8% of all early pregnancy samples) in our cohort may have limited our power to detect such associations. Low prevalence of *Lactobacillus* depleted vaginal communities is likely attributable to increased relative abundance of *Lactobacillus* species in the vaginal microbiome during pregnancy and consistent with our clinical population: mostly women with European ancestry who, compared to other ethnicities, have higher prevalence of *Lactobacillus*-dominated vaginal microbiomes ^28,30^. A recent study of 60 women presenting with suspected preterm labour reported positive associations between specific urinary and blood human milk oligosaccharides, and some vaginal taxa and clinical outcomes including preterm birth ^31^. However, it is difficult to compare these findings to our own given that over half of their patient cohort received tocolysis to inhibit uterine contractions and prevent preterm birth (only a small number of women in their study cohort experienced preterm birth, n=11).

To investigate whether vaginal microbial community structure might differ between secretors and non-secretors, we compared co-occurrence networks inferred from correlation matrices based on the 16S relative abundance data (Figure 2) as previously described ^32^. For this analysis, non-*Lactobacillus* taxa were classified as ‘pathobionts’, ‘BV-associated’ or ‘other’ using definitions from Wijgert et al ^33^. In both the secretor and non-secretor microbial networks, nodes representing BV-associated bacteria tended to be connected via positive edges, reflecting positive correlations among these microbes (Figure 2A). Analysis of node importance, using Expected Influence (EI) ^34^ as a metric, confirmed this pattern, i.e., that in both networks, nodes with the most positive EI scores were BV-associated bacteria and pathobionts, such as *Prevotella timonensis* and *Finegoldia magna* (Supplementary Figure S2). These patterns mirror previous studies showing that these microbes establish interactions based on mutualistic relationships ^35,36^. In non-secretors *L. crispatus, L. jensenii* and *L. iners* were the most negatively scored nodes for EI, but in secretors, a greater proportion of positive edges from these lactobacilli to other BV-associated microbes was observed (Figure 2B and Supplementary Figure S2). These results suggest that *Lactobacillus* species, particularly *L. crispatus*, are more refractory to co-colonisation in non-secretors where they may offer a greater “protective” effect via competitive exclusion. These findings are consistent with comparative genomics studies that have highlighted bacterial cell surface glycoconjugates and carbohydrate-binding proteins as potentially important mediators of vaginal microbiota-host ^37^.

**Figure 2:**
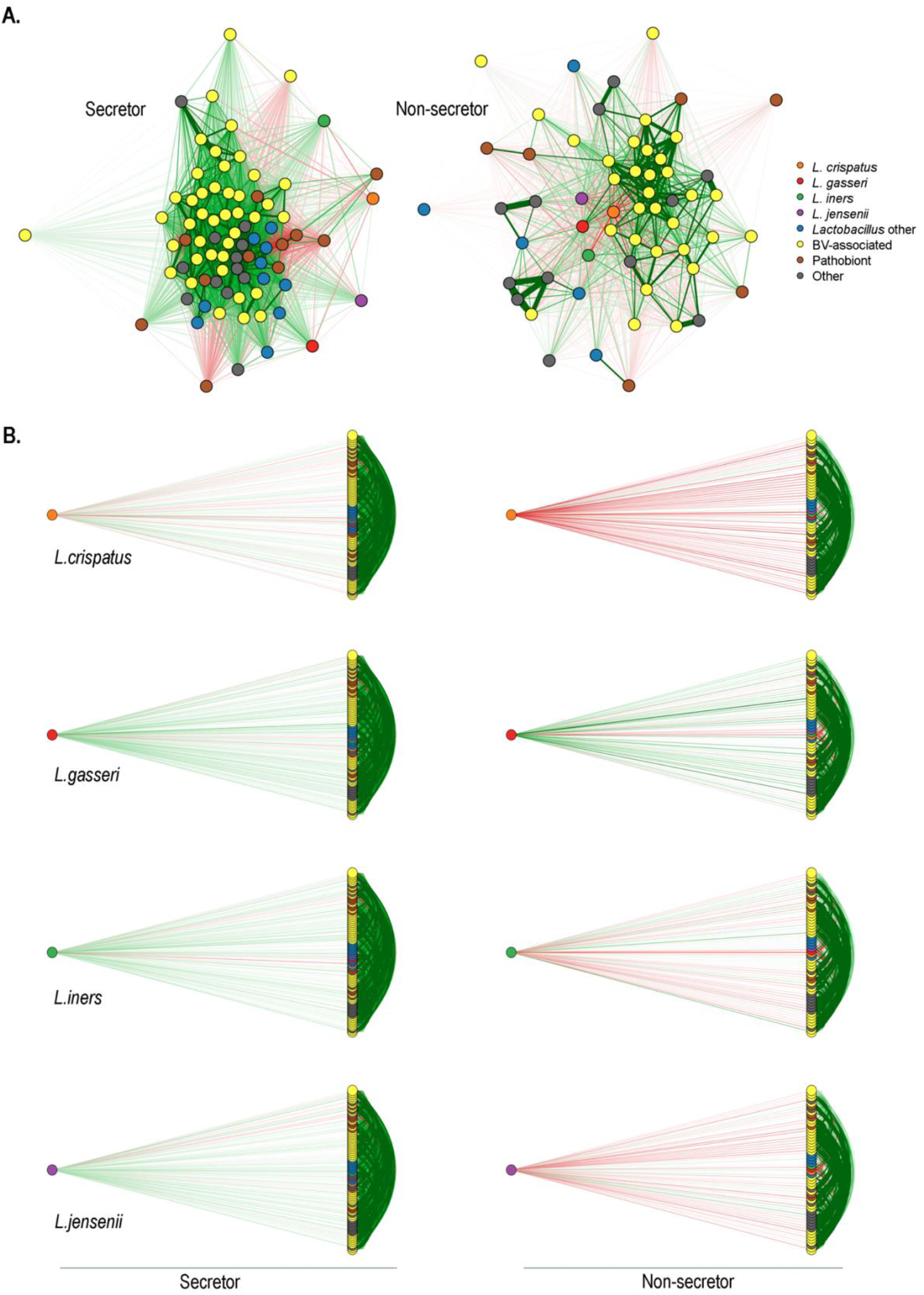
A. Co-occurrence networks showing the vaginal microbiata in early pregnancy in secretors (left) and non-secretors (right). Nodes represent bacterial taxa present in the 16S relative abundance data (above our low abundance threshold of 0.5% in two or more samples), and have been grouped into *L. crispatus, L. gasseri, L. iners, L. jensenii*, other *Lactobacillus*, BV-associated bacteria, Pathobionts, and Other, as defined by previously ^33^. Statistically significant edges, indicating correlations, are coloured and weighted according to their r value: red and green, respectively indicate negatively and positively correlations. In both secretors and non-secretors, many of the positive associations occur between BV-associated microbes, reflecting the typically polymicrobial nature of BV. B. Flow diagrams (derived from the network analyses) centred on the *L. crispatus, L. gasseri, L. iners* and *L. jensenii* nodes indicate that these tend to be more negatively correlated with other bacteria in the vaginal microbiota of the non-secretors compared with the secretors with *L. crispatus* having the greatest proportion of negative edges, particularly in the non-secretors.

### Non-secretors with Lactobacillus depleted microbiomes are associated with shorter gestational length

On the basis of the distribution of *Lactobacillus* abundance (Figure 1C) and previous studies ^38,39^ we classified the vaginal microbiome into *Lactobacillus* dominated (>90% *Lactobacillus* spp.) and depleted states (<90%). Non-secretors with a *Lactobacillus* depleted microbiome in early pregnancy had a shorter mean gestational length 245.5 days (SD=44.5) compared to *Lactobacillus* dominated non-secretors, 265.9 days (SD=23.6) and to both *Lactobacillus* depleted and dominated secretors, 264.3 (SD=21.2) and 261.8 (SD=27.5) days, respectively (Figure 3A). However, the association between *Lactobacillus* status in non-secretors on gestational outcome was diminished by mid pregnancy. To confirm these observations, for each sampling timepoint we modelled gestational length on secretor status and the vaginal microbiome (*Lactobacillus* status) together with BMI, age, cervical cerclage status and previous pregnancy history, i.e. cervical excisional treatment (LLETZ) and a combined previous preterm birth (PTB) and/or mid-trimester loss (MTL) covariate using generalised linear mixed effects modelling (GLMM) with ethnicity as a random effect (Table 1). As expected, previous pregnancy histories were significant explanatory variables of gestational length (Supplementary Table S3). Comparison of the GLMMs with generalised linear models (GLM) showed similar model fits (for example, AIC 2495.225 and 2497.225 for the GLM and GLMM, respectively, for the early pregnancy modelling) indicating that ethnicity was not a significant factor in the model. However, the early pregnancy GLMM showed that the secretor status and *Lactobacillus* status interaction term is a significant explanatory variable of gestational length (p=0.02). This significance disappeared by mid-pregnancy supporting our earlier observation that non-secretors with a *Lactobacillus* depleted vaginal microbiome in early pregnancy, but not mid-pregnancy, are associated with shorter gestational lengths. *Post hoc* testing using the estimated marginal means confirmed this significant comparison (p-value=0.045), but also indicated that a difference in gestational length may exist between *Lactobacillus* depleted non-secretors (mean gestation of 245.5 days) and *Lactobacillus* depleted secretors (mean gestation of 264.3 days) (p-value=0.068). No significant difference between *Lactobacillus* dominated and depleted secretors was found (p-value=0.283).

**Table 1.** Unstandardised coefficients (*b*), standard error (SE) and 95% confidence intervals (CI) from gamma generalised linear mixed effects modelling (GLMM) of gestational length (in days) over three time points in pregnancy (with ethnicity as a random effect). Early, Mid and Late pregnancy GLMMs using *Lactobacillus* status (dominated/depleted), as well as an Early Community State Type (CST) GLMM, which replaces the *Lactobacillus* covariate with CST. Secretors with *Lactobacillus* dominated microbiota (or CST 1) are baseline in the model. *^1^ Preterm birth. *^2^ Mid-trimester loss. * p<0.05. ** p<0.01. *** p<0.001.

|  | Regression model |  |  |  |  |  |  |  |
| --- | --- | --- | --- | --- | --- | --- | --- | --- |
|  | Early<br>b (SE) | CI | Mid<br>b (SE) | CI | Late<br>b (SE) | CI | Early (CST)<br>b (SE) | CI |
| Intercept | 3.16 (0.28) | 2.62,3.71 | 3.04 (0.29) | 2.48,3.60 | 2.91 (0.29) | 2.35,3.48 | 3.15 (0.29) | 2.58,3.72 |
| Age | 0.002 (0.01) | -0.01,0.02 | 0.01 (0.01) | -0.01,0.02 | 0.01 (0.08) | -0.01,0.03 | 0.01 (0.01) | -0.01,0.02 |
| BMI (<18.5) | -0.12 (0.41) | -0.92,0.68 | -0.01 (0.34) | -0.68,0.67 | 0.02 (0.38) | -0.72,0.75 | -0.1 (0.41) | -0.91,0.71 |
| BMI (25.0-29.99) | 0.09 (0.09) | -0.10,0.27 | 0.01 (0.1) | -0.18,0.19 | 0.09 (0.1) | -0.10,0.28 | 0.1 (0.1) | -0.09,0.29 |
| BMI (>30.0) | -0.16 (0.13) | -0.41,0.09 | -0.05 (0.12) | -0.28,0.18 | -0.13 (0.12) | -0.37,0.11 | -0.21 (0.13) | -0.47,0.05 |
| Cervical stitch | 0.33 (0.09)*** | 0.16,0.50 | 0.34 (0.08)*** | 0.17,0.50 | 0.21 (0.08)* | 0.04,0.37 | 0.32 (0.1)*** | 0.15,0.49 |
| Previous<br>PTB*<sup>1</sup>/MTL*<sup>2</sup> | 0.35 (0.1)*** | 0.16,0.54 | 0.31 (0.1)** | 0.12,0.50 | 0.23 (0.1)* | 0.04,0.43 | 0.35 (0.1)*** | 0.15,0.54 |
| Previous<br>cervical<br>excisional<br>treatment | -0.28 (0.1)** | -0.48,-0.08 | -0.28 (0.1)** | -0.47,-0.08 | -0.26 (0.1)* | -0.45,-0.06 | -0.31 (0.1)** | -0.52,-0.11 |
| Secretor ×<br>*Lactobacillus* | 0.53 (0.23)* | 0.07,0.99 | 0.2 (0.24) | -0.27,0.67 | -0.16 (0.23) | -0.61,0.29 | NA | NA |
| Secretor ×<br>CST 2 | NA | NA | NA | NA | NA | NA | 0.84 (0.34)* | 0.17,1.51 |
| Secretor ×<br>CST 3 | NA | NA | NA | NA | NA | NA | 0.43 (0.22)* | 0.00,0.86 |
| Secreter ×<br>CST 4 | NA | NA | NA | NA | NA | NA | 0.85 (0.35)* | 0.17,1.52 |
| Secreter ×<br>CST 5 | NA | NA | NA | NA | NA | NA | -0.11 (0.43) | -0.95,0.73 |

**Figure 3:**
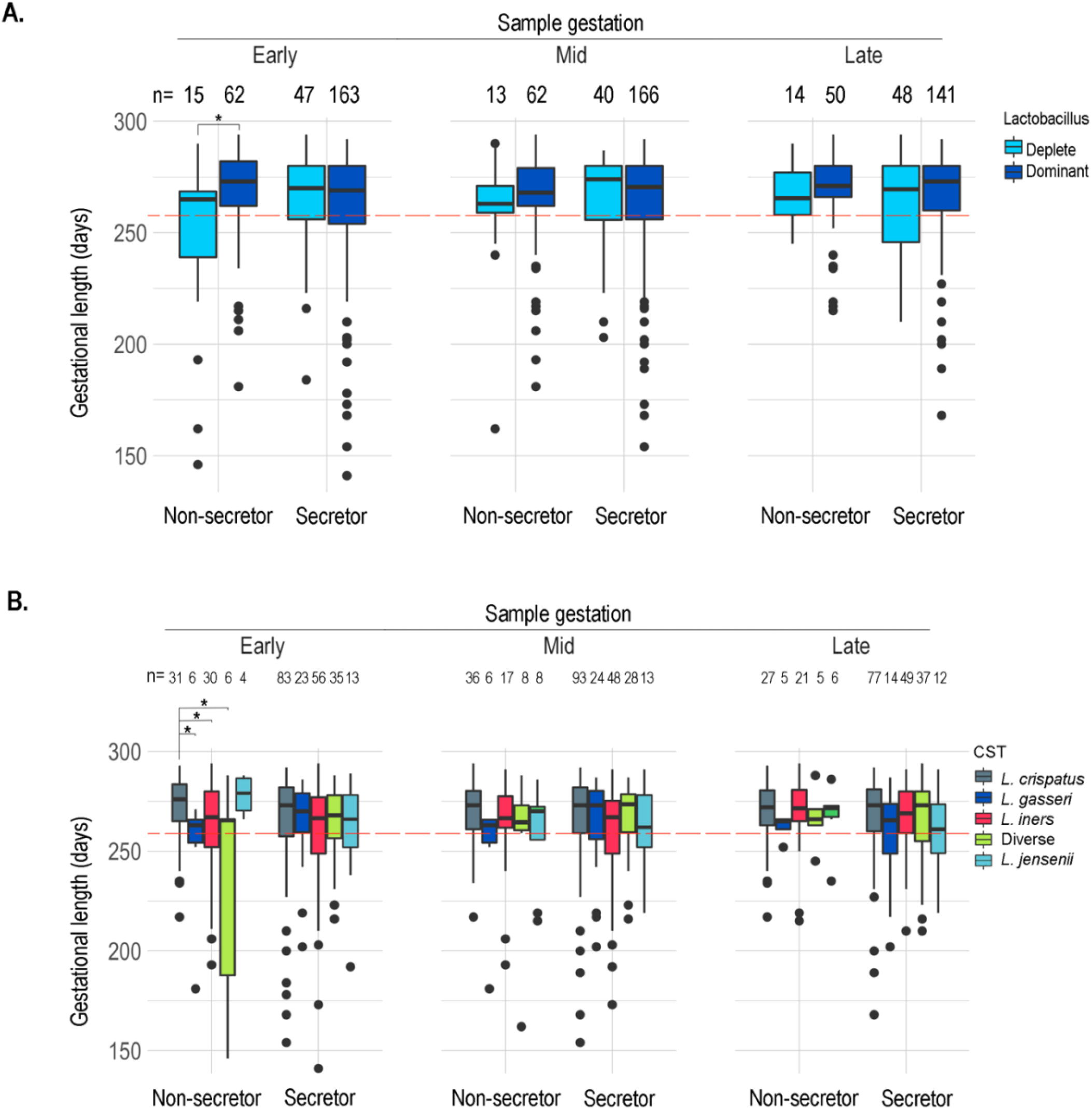
The association between secretor status, the vaginal microbiome and gestational length (days). A red dashed line highlights the 37 weeks (259 days) threshold for prematurity and sample sizes are indicated at the top of the plots. A. The distribution of gestational length by secretor status and vaginal microbiome shows non-secretors with *Lactobacillus* deplete microbiomes in early pregnancy tend to have shorter gestation but this relationship disappears by mid pregnancy. B. Classifying the microbiome by Community State Type (CST) rather than by *Lactobacillus* status indicates that reduced gestation in *Lactobacillus* deplete non-secretors is mainly driven by women with CST 2 and high diversity CST 4 microbiomes.

When vaginal microbiomes were classified into CSTs, non-secretors with CST 2, 3 or 4 in early pregnancy had shorter gestations, mean (±SD) of 249.3 (34.1), 260.5 (26.9) and 232.2 (61.4) days, respectively, compared to CST 1 (269.4 days (19.7)) (Figure 3B). A similar pattern was observed with CST 5 (mean of 278 days), however sample size was limited to only n=4. By contrast, gestational length in secretors was broadly similar across all CSTs: mean gestational length (±SD) of 264.0 (28.4), 264.7 (20.5), 258.0 (29.4), 264.1 (18.2) and 262.6 (26.0) respectively in CST 1, 2, 3, 4 and 5. In line with these findings, modelling gestational length with early pregnancy CST (Table 1) showed that the interaction term between CST and secretor status was statistically significant (p-value=0.017). By mid-pregnancy, this interaction term was not statistically significant (Supplementary Table S4). The estimated marginal means identified the gestational length differences in the non-secretors between CST 1 versus CST 2, and CST 1 versus CST 4 to be statistically significant (respectively, p-value=0.037 and 0.033). Trend differences were also observed in the non-secretors between CST 2 and 5 (p-value=0.059), and CST 4 and 5 (p-value=0.051). These *post hoc* tests also identified a difference in gestational length between secretors and non-secretors with CST 1 vaginal microbiomes in early pregnancy (p-value=0.017).

Our findings are consistent with a previous study by Lurie *et al* ^40^ who identified a significantly elevated proportion of non-secretors in a cohort of 28 patients who suffered preterm premature rupture of the membranes (PPROM) compared with term patients. PPROM precedes approximately 30% of all preterm birth cases and has been associated with ascending infection in the vagina ^18,41^.

Caldwell et al ^42^ also reported that maternal secretor status is a potential biomarker for prematurity. Our data supports these findings but demonstrates that this relationship is likely shaped by the vaginal microbiota in early pregnancy. Dominance of the vaginal niche by *Lactobacillus* spp. is considered to be a hallmark of vaginal health due to the role that these species play in preventing colonisation of other microbes through the promotion of a hostile, acidic mucosal environment enriched by bacteriocins and other antimicrobial compounds ^43,44^. Our results support previous findings in highlighting this protective role extending into pregnancy, but demonstrate that this is nuanced by host secretor status. Studies identifying an association between vaginal *Lactobacillus* spp. depletion and increased risk of preterm birth ^19,28,45,46^ also emphasise that high diversity vaginal bacterial communities do not always result in preterm birth (as is the case of the secretors). An enhanced inflammatory response, previously observed in non-secretors ^47^, may provide a mechanism linking gestational length to secretor status and vaginal microbiota but this requires further investigation. It is possible that ABO blood groups influence the relationship between secretor status and the microbiota by affecting terminal glycan structures. To address this, we incorporated ABO status into our models and observed that there may be evidence of shorter gestational length in blood group B women compared to blood group A, though this was only observed at mid-pregnancy (Supplementary Table S5).

Early developmental stages in pregnancy have lasting effects on pregnancy gestation and outcome. Our results indicate that the relationship between *Lactobacillus* depletion and reduced gestational length in non-secretors is altered as pregnancy progresses and points towards the microbiota-host interactions in early pregnancy being particularly key in shaping preterm birth risk. This result is consistent with findings from Stout and colleagues who showed that, in a predominantly African-American cohort, early gestation is an ecologically important time for events that predict subsequent term and preterm birth ^48^. Similarly, Tabatabaei and coworkers ^49^ identified an association between *Lactobacillus* depleted vaginal microbiota in early pregnancy and increased risk of early preterm birth in a cohort of Canadian women of predominantly white European origin. *Lactobacillus* depletion is also a risk factor for miscarriage, which collectively suggests a potential effect of the vaginal microbiota upon decidual function and placentation ^17,50,51^.

### Concluding remarks

In this study we identify a subset of women, non-secretors with *Lactobacillus* depleted vaginal microbiomes, who are predisposed to shorter gestational length. This stratification could allow a more targeted intervention of “at-risk” pregnancies, although these results should be verified with larger numbers of *Lactobacillus* depleted samples. We find evidence of a more “protective” role of *Lactobacillus* species, especially *L. crispatus*, in non-secretors. Whilst further studies are required to elucidate the mechanisms underlying these interactions in secretors and non-secretors, the data supports the evolving concept that *L. crispatus* offers optimal protection against preterm birth and is a candidate for development of live biotherapeutic therapies during pregnancy.

## Data Availability

All sequencing data used in this study has been made available in ENA database (Accession numbers ERR4864561 to ERR4865307). Python and R scripts used in support of the analyses are available at: https://github.com/samitkundu/FUT2. Further information and requests for supporting data, resources, and reagents should be directed to the Lead Contact: David MacIntyre.

https://www.ebi.ac.uk/ena/browser/view/ERR4864561-ERR4865307

https://github.com/samitkundu/FUT2

## Resource Availability

Further information and requests for supporting data, resources, and reagents should be directed to the Lead Contact: David MacIntyre.

### Materials Availability

Reagents from this study are available upon request.

### Data and Code Availability

All sequencing data used in this study has been made available in ENA database (Accession numbers ERR4864561-ERR4865307). Python and R scripts used in support of the analyses are available at: https://github.com/samitkundu/FUT2.

## Acknowledgements

The study was funded by the March of Dimes. The funder had no role in the study design, data collection and analysis, and preparation of the manuscript.

## Author Contributions

Conceptualization, D.A.M., P.R.B. and S.K.; Methodology, D.A.M., P.R.B. and S.K.; Investigation, D.C., R.G.B., L.K., S.K. and Y.S.L.; Resources, D.C., H.L., L.K., L.S. and R.G.B.; Formal Analysis, S.K.; Writing – Original draft, D.A.M., P.R.B. and S.K.; Writing – Review and Editing, A.D., D.A.M., H.L., J.M., L.S., P.R.B., S.H., S.K., T.F., Y.L., Y.S.L.; Funding Acquisition, D.A.M. and P.R.B.; Supervision, D.A.M. and P.R.B.

## Methods

### Ethics Statement

The study was conducted with approval of the NHS National Research Ethics Service (NRES) Committees London -City and East (REC 12/LO/2003) and London– Stanmore (REC 14/LO/0328), and by the North of Scotland Research Ethics Service (REC 14/NS/1078). Written informed consent was obtained from all patients prior to sampling and experiments were performed in accordance with the approved institutional guidelines.

### Patient sampling

Recruitment and sampling were performed at Imperial College Healthcare NHS Trust Hospitals (Queen Charlotte’s and Chelsea and St Mary’s Hospitals), London, UK, at Chelsea & Westminster Hospital (NHS Trust, London, UK), University College London Hospital (NHS Foundation Trust, London, UK) and the Royal Infirmary of Edinburgh, Scotland, UK. Eligibility criteria included singleton pregnancies, with and without risk factors for preterm birth. Exclusion criteria included women under 18 years of age, those who had sexual intercourse within 72 h of sampling, vaginal bleeding in the preceding week, antibiotic use in the preceding 2 weeks, multiple pregnancies, HIV or Hepatitis C positive status. Detailed maternal clinical metadata and birth outcome data was collected for all participants. Cervicovaginal fluid swab samples were collected from the posterior fornix using BBL CultureSwab MaxV Liquid Amies swabs (Becton, Dickinson and Company, Oxford, UK) at up to three timepoints throughout pregnancy; early (63 to 130 days), mid (118-180 days) and late (163-252 days). Swabs were immediately placed in Amies transport media and stored at -80°C.

### Determination of secretor status

Extraction of DNA from vaginal swabs was performed as previously described ^30^. Individuals were genotyped by sequencing the coding part of exon 2 of the *FUT2* gene. The exon was amplified using primers 5’-CCATATCCCAGCTAACGTGTCC-3’ and 5’-GGGAGGCAGAGAAGGAGAAAAGG-3 ^52^ and the amplicons sequenced with the PacBio Sequel system. Primer sequences were removed from the CCS reads using Dada2 ^53^ and the trimmed sequences mapped to a human reference FUT2 sequence (derived from HG38) using Minimap ^54^. To limit the effect of any reference bias, we generated consensus sequences using the “ALT” allele from the mapping and reads were remapped to this sequence. Finally, variants were called using Freebayes ^55^ and phased using WhatsHap ^56^.

Individuals that were homozygous for any one of the four known non-secretor mutations (se302 (P101L, rs200157007), se385 (I129F, rs1047781), se428 (W413X, rs601338) and se571(R191X, rs18000028)) were inferred to be phenotypically non-secretor ^4–6,57^. Heterozygotes and wildtype homozygotes were classified as secretors ^27^.

### Sequencing of 16S rRNA gene amplicons and assembly

The V1-V2 hyper variable regions of bacterial 16S rRNA genes were amplified with using the forward primer set (28f-YM) consisting of a mixture of the following primers mixed at a 4:1:1:1 ratio; 28F-Borrellia GAGTTTGATCCTGGCTTAG; 28F-Chlorflex GAATTTGATCTTGGTTCAG; 28F-Bifido GGGTTCGATTCTGGCTCAG; 28F GAGTTTGATCNTGGCTCAG. The reverse primer consisted of; 388R TGCTGCCTCCCGTAGGAGT ^58^. Primer sequences were trimmed using Cutadapt ^59^, performed QC using FastQC ^60^ and calculated ASV counts per sample using the Qiime2 pipeline ^61^. We used DADA2 ^53^ for denoising and taxonomically classified sequences to species level using the STIRRUPS reference database ^62^.

### Data analyses

Generalised linear mixed effects gamma regression models for each timepoint in pregnancy were used to regress gestational length (in days) on secretor status and the vaginal microbiome (*Lactobacillus* status or CST) as well as age, BMI, previous PTB/MTL, cervical stitch and Previous cervical excisional treatment in glmmTMB ^63^. We used ethnicity as a random effect: as the sample size for the East Asian and Middle East North African and Mixed/Other groups was small (less than 10) these were combined into a single group (Mixed/Other). We applied a reflection transformation to the gestational length prior to fitting the gamma GLMM as this variable was negatively skewed. Model diagnostics were examined with scaled residuals simulated from the fitted models using DHARMa ^64^. We used the emmeans package to calculate the estimated marginal means from the models to look for statistically significant comparisons between groups ^65^: this *post hoc* method automatically adjusts for multiple comparisons.

We retained taxa that occurred at greater than 0.5% abundance in two or more samples. Differential abundance analyses were performed using ALDEx2 to compare taxon abundances in the vaginal microbiome in early pregnancy between secretors and non-secretors ^66,67^. This method uses a compositionally robust approach to perform DAA: microbiome count data are considered compositional and thus have to be transformed to permit analysis by multivariate statistics ^68^. Count data were transformed with the centred log-ratio (CLR) and we ran 1000 Monte-Carlo instances to estimate effect sizes and perform a Welch’s t test to compare our two conditions. We also analysed the compositional differences between phenotypes using PCAs: zeros in the count data were imputed using a Bayesian multiplicative approach in the zCompositions package ^69^ followed by CLR-transformation. To test whether the centroids of the two secretor groups differ we performed a PERMANOVA on the CLR-transformed distance matrix (after checking for homogeneity of dispersion) using the Vegan package ^70^.

Finally, we used BAnOCC to infer co-occurrence networks for the microbial taxa observed in secretors and non-secretors in early pregnancy ^71^. This program uses a Bayesian framework to analyse compositional covariance. We ran the MCMC for 10000 generations and four chains to reach convergence. We used a 95% credible interval to identify significant correlations. Qgraph was used to draw the networks based on these Banocc-derived adjacency matrices and to calculate the Expected Influence network statistic, which is a measure of degree centrality for signed networks^34,72^.

## References

1. Audfray A, Varrot A, Imberty A. Bacteria love our sugars: Interaction between soluble lectins and human fucosylated glycans, structures, thermodynamics and design of competing glycocompounds. Comptes Rendus Chim. 2013 May;16(5):482–90.

2. Marcobal A, Southwick AM, Earle KA, Sonnenburg JL. A refined palate: Bacterial consumption of host glycans in the gut. Glycobiology. 2013 Sep 1;23(9):1038–46.

3. McGuckin MA, Lindén SK, Sutton P, Florin TH. Mucin dynamics and enteric pathogens. Nat Rev Microbiol. 2011;9(9):265–78.

4. Kelly RJ, Rouquier S, Giorgi D, Lennon GG, Lowe KB. Sequence and expression of a candidate for the Human Secretor blood group alpha(1,2)Fucosyltransferase gene (FUT2). J Biol Chem. 1995;270(270):4640–9.

5. Soejima M, Pang H, Koda Y. Genetic variation of FUT2 in a Ghanaian population: identification of four novel mutations and inference of balancing selection. Ann Hematol. 2007 Jan 26;86(3):199–204.

6. Birney E, Stamatoyannopoulos JA, Dutta A, Guigó R, Gingeras TR, Margulies EH, et al. Identification and analysis of functional elements in 1% of the human genome by the ENCODE pilot project. Nature. 2007;447(447):799–816.

7. Ferrer-Admetlla A, Sikora M, Laayouni H, Esteve A, Roubinet F, Blancher A, et al. A natural history of FUT2 polymorphism in humans. Mol Biol Evol. 2009;26(26):1993–2003.

8. Carlsson B, Kindberg E, Buesa J, Rydell GE, Lidón MF, Montava R, et al. The G428A Nonsense Mutation in FUT2 Provides Strong but Not Absolute Protection against Symptomatic GII.4 Norovirus Infection. Lopman BA, editor. PLoS One. 2009 May 18;4(5):e5593.

9. Blackwell CC, Jónsdóttir K, Hanson M, Todd WTA, Chaudhuri AKR, Mathew B, et al. Non-secretion of ABO antigens predisposing to infection by Neisseria meningitidis and Streptococcus pneumoniae. Lancet. 1986 Aug;328(8501):284–5.

10. Blackwell CC, Jonsdottir K, Hanson MF, Weir DM. Non-secretion of ABO blood group antigens predisposing to infection by Haemophilus influenzae. Lancet. 1986 Sep;328(8508):687.

11. Wacklin P, Tuimala J, Nikkilä J, Sebastian Tims, Mäkivuokko H, Alakulppi N, et al. Faecal Microbiota Composition in Adults Is Associated with the FUT2 Gene Determining the Secretor Status. Quince C, editor. PLoS One. 2014 Apr 14;9(4):e94863.

12. Rausch P, Rehman A, Kunzel S, Hasler R, Ott SJ, Schreiber S, et al. Colonic mucosa-associated microbiota is influenced by an interaction of Crohn disease and FUT2 (Secretor) genotype. Proc Natl Acad Sci. 2011 Nov 22;108(47):19030–5.

13. Wacklin P, Mäkivuokko H, Alakulppi N, Nikkilä J, Tenkanen H, Räbinä J, et al. Secretor genotype (FUT2 gene) is strongly associated with the composition of bifidobacteria in the human intestine. PLoS One. 2011;6(5).

14. Lewis ZT, Totten SM, Smilowitz JT, Popovic M, Parker E, Lemay DG, et al. Maternal fucosyltransferase 2 status affects the gut bifidobacterial communities of breastfed infants. Microbiome. 2015 Dec 10;3(1):13.

15. Bayar E, Bennett PR, Chan D, Sykes L, MacIntyre DA. The pregnancy microbiome and preterm birth. Semin Immunopathol. 2020 Aug 14;42(4):487–99.

16. Bennett PR, Brown RG, MacIntyre DA. Vaginal Microbiome in Preterm Rupture of Membranes. Obstet Gynecol Clin North Am. 2020 Dec;47(4):503–21.

17. Al Memar M, Bobdiwala S, Fourie H, Mannino R, Lee Y, Smith A, et al. The association between vaginal bacterial composition and miscarriage: a nested case–control study. BJOG An Int J Obstet Gynaecol. 2020 Jan 31;127(2):264–74.

18. Brown RG, Marchesi JR, Lee YS, Smith A, Lehne B, Kindinger LM, et al. Vaginal dysbiosis increases risk of preterm fetal membrane rupture, neonatal sepsis and is exacerbated by erythromycin. BMC Med. 2018;16(16):1–15.

19. Callahan BJ, DiGiulio DB, Goltsman DSA, Sun CL, Costello EK, Jeganathan P, et al. Replication and refinement of a vaginal microbial signature of preterm birth in two racially distinct cohorts of US women. Proc Natl Acad Sci. 2017;114(114):9966–71.

20. Pike KC, Lucas JSA. Respiratory consequences of late preterm birth. Paediatr Respir Rev. 2015;16(16):182–8.

21. Lax ID, Duerden EG, Lin SY, Mallar Chakravarty M, Donner EJ, Lerch JP, et al. Neuroanatomical consequences of very preterm birth in middle childhood. Brain Struct Funct. 2013;218(218):575–85.

22. Chehade H, Simeoni U, Guignard J-P, Boubred F. Preterm Birth: Long Term Cardiovascular and Renal Consequences. Curr Pediatr Rev. 2018;14(14):219–26.

23. Goldenberg RL, Culhane JF, Iams JD, Romero R. Epidemiology and causes of preterm birth. Lancet. 2008 Jan;371(9606):75–84.

24. Kyrgiou M, Mitra A, Arbyn M, Stasinou SM, Martin-Hirsch P, Bennett P, et al. Fertility and early pregnancy outcomes after treatment for cervical intraepithelial neoplasia: systematic review and meta-analysis. BMJ. 2014 Oct 28;349(oct28 1):g6192–g6192.

25. Mitra A, MacIntyre DA, Lee YS, Smith A, Marchesi JR, Lehne B, et al. Cervical intraepithelial neoplasia disease progression is associated with increased vaginal microbiome diversity. Sci Rep. 2015 Dec 17;5(1):16865.

26. Mitra A, MacIntyre DA, Ntritsos G, Smith A, Tsilidis KK, Marchesi JR, et al. The vaginal microbiota associates with the regression of untreated cervical intraepithelial neoplasia 2 lesions. Nat Commun. 2020 Dec 24;11(1):1999.

27. Fumagalli M, Cagliani R, Pozzoli U, Riva S, Comi GP, Menozzi G, et al. Widespread balancing selection and pathogen-driven selection at blood group antigen genes. Genome Res. 2009;19(19):199–212.

28. Fettweis JM, Serrano MG, Brooks JP, Edwards DJ, Girerd PH, Parikh HI, et al. The vaginal microbiome and preterm birth. Nat Med. 2019;25(25):1012–21.

29. Brooks JP, Buck GA, Chen G, Diao L, Edwards DJ, Fettweis JM, et al. Changes in vaginal community state types reflect major shifts in the microbiome. Microb Ecol Health Dis. 2017 Jan 1;28(1):1303265.

30. MacIntyre DA, Chandiramani M, Lee YS, Kindinger L, Smith A, Angelopoulos N, et al. The vaginal microbiome during pregnancy and the postpartum period in a European population. Sci Rep. 2015;5(5):8988.

31. Pausan M-R, Kolovetsiou-Kreiner V, Richter GL, Madl T, Giselbrecht E, Obermayer-Pietsch B, et al. Human Milk Oligosaccharides Modulate the Risk for Preterm Birth in a Microbiome-Dependent and -Independent Manner. Jansson JK, editor. mSystems. 2020 Jun 30;5(3).

32. Ma B, Wang Y, Ye S, Liu S, Stirling E, Gilbert JA, et al. Earth microbial cooccurrence network reveals interconnection pattern across microbiomes. Microbiome. 2020 Dec 4;8(1):82.

33. van de Wijgert JHHM, Verwijs MC, Gill AC, Borgdorff H, van der Veer C, Mayaud P. Pathobionts in the Vaginal Microbiota: Individual Participant Data Meta-Analysis of Three Sequencing Studies. Front Cell Infect Microbiol. 2020;10(April).

34. Robinaugh DJ, Millner AJ, McNally RJ. Identifying highly influential nodes in the complicated grief network. J Abnorm Psychol. 2016;125(125):747–57.

35. Pybus V, Onderdonk AB. Evidence for a Commensal, Symbiotic Relationship between Gardnerella vaginalis and Prevotella bivia Involving Ammonia: Potential Significance for Bacterial Vaginosis. J Infect Dis. 1997 Feb 1;175(2):406–13.

36. Machado A, Jefferson K, Cerca N. Interactions between Lactobacillus crispatus and Bacterial Vaginosis (BV)-Associated Bacterial Species in Initial Attachment and Biofilm Formation. Int J Mol Sci. 2013 Jun 5;14(6):12004–12.

37. van der Veer C, Hertzberger RY, Bruisten SM, Tytgat HLP, Swanenburg J, de Kat Angelino-Bart A, et al. Comparative genomics of human Lactobacillus crispatus isolates reveals genes for glycosylation and glycogen degradation: implications for in vivo dominance of the vaginal microbiota. Microbiome. 2019;7(7):49.

38. Moreno I, Codoñer FM, Vilella F, Valbuena D, Martinez-Blanch JF, Jimenez-Almazán J, et al. Evidence that the endometrial microbiota has an effect on implantation success or failure. Am J Obstet Gynecol. 2016 Dec;215(6):684–703.

39. Fu M, Zhang X, Liang Y, Lin S, Qian W, Fan S. Alterations in Vaginal Microbiota and Associated Metabolome in Women with Recurrent Implantation Failure. Fidel PL, Lin X, editors. MBio. 2020 Jun 2;11(3).

40. Lurie S, Ben-Aroya Z, Eldar S, Sadan O. Association of Lewis blood group phenotype with preterm premature rupture of membranes. J Soc Gynecol Investig. 2003;10(10):291–3.

41. Benedetto C, Tibaldi C, Marozio L, Marini S, Masuelli G, Pelissetto S, et al. Cervicovaginal infections during pregnancy: epidemiological and microbiological aspects. J Matern Neonatal Med. 2004 Aug 1;16(2):9–12.

42. Caldwell J, Matson A, Mosha M, Hagadorn JI, Moore J, Brownell E. Maternal H-antigen secretor status is an early biomarker for potential preterm delivery. J Perinatol. 2020 Nov 24;

43. Tamrakar R, Yamada T, Furuta I, Cho K, Morikawa M, Yamada H, et al. Association between Lactobacillus species and bacterial vaginosis-related bacteria, and bacterial vaginosis scores in pregnant Japanese women. BMC Infect Dis. 2007;7:128.

44. MacIntyre DA, Sykes L, Bennett PR. The human female urogenital microbiome: complexity in normality. Marchesi JR, editor. Emerg Top Life Sci. 2017 Nov 30;1(4):363–72.

45. Elovitz MA, Gajer P, Riis V, Brown AG, Humphrys MS, Holm JB, et al. Cervicovaginal microbiota and local immune response modulate the risk of spontaneous preterm delivery. Nat Commun. 2019;10(10):1–8.

46. Brown RG, Al-Memar M, Marchesi JR, Lee YS, Smith A, Chan D, et al. Establishment of vaginal microbiota composition in early pregnancy and its association with subsequent preterm prelabor rupture of the fetal membranes. Transl Res. 2019 May;207:30–43.

47. Lomberg H, Jodal U, Leffler H, Man P De, Svanborg C. Blood Group Non-Secretors Have an Increased Inflammatory Response to Urinary Tract Infection. Scand J Infect Dis. 1992 Jan 8;24(1):77–83.

48. Stout MJ, Zhou Y, Wylie KM, Tarr PI, Macones GA, Tuuli MG. Early pregnancy vaginal microbiome trends and preterm birth. Am J Obstet Gynecol. 2017 Sep;217(3):356.e1-356.e18.

49. Tabatabaei N, Eren AM, Barreiro LB, Yotova V, Dumaine A, Allard C, et al. Vaginal microbiome in early pregnancy and subsequent risk of spontaneous preterm birth: a case–control study. BJOG An Int J Obstet Gynaecol. 2019;126(126):349–58.

50. Kirkegaard I, Uldbjerg N, Petersen OB, Tørring N, Henriksen TB. PAPP-A, free β-hCG, and early fetal growth identify two pathways leading to preterm delivery. Prenat Diagn. 2010 Aug 18;30(10):956–63.

51. Lackman F, Capewell V, Richardson B, DaSilva O, Gagnon R. The risks of spontaneous preterm delivery and perinatal mortality in relation to size at birth according to fetal versus neonatal growth standards. Am J Obstet Gynecol. 2001 Apr;184(5):946–53.

52. Silva LM, Carvalho AS, Guillon P, Seixas S, Azevedo M, Almeida R, et al. Infection-associated FUT2 (Fucosyltransferase 2) genetic variation and impact on functionality assessed by in vivo studies. Glycoconj J. 2010 Jan 16;27(1):61–8.

53. Callahan BJ, McMurdie PJ, Rosen MJ, Han AW, Johnson AJA, Holmes SP. DADA2: High-resolution sample inference from Illumina amplicon data. Nat Methods. 2016 Jul 23;13(7):581–3.

54. Li H. Minimap2: pairwise alignment for nucleotide sequences. Birol I, editor. Bioinformatics. 2018 Sep 15;34(18):3094–100.

55. Garrison E, Marth G. Haplotype-based variant detection from short-read sequencing. 2012 Jul 17;

56. Patterson M, Marschall T, Pisanti N, van Iersel L, Stougie L, Klau GW, et al. WhatsHap : Weighted Haplotype Assembly for Future-Generation Sequencing Reads. J Comput Biol. 2015 Jun;22(6):498–509.

57. Henry S, Mollicone R, Fernandez P, Samuelsson B, Oriol R, Larson G. Molecular basis for erythrocyte Le(a+b+) and salivary ABH partial-secretor phenotypes: expression of a FUT2 secretor allele with an A-T mutation at nucleotide 385 correlates with reduced a(1,2) fucosyltransferase activity. Glycoconj J. 1996 Dec;13(6):985–93.

58. Frank JA, Reich CI, Sharma S, Weisbaum JS, Wilson BA, Olsen GJ. Critical Evaluation of Two Primers Commonly Used for Amplification of Bacterial 16S rRNA Genes. Appl Environ Microbiol. 2008 Apr 15;74(8):2461–70.

59. Martin M. Cutadapt removes adapter sequences from high-throughput sequencing reads. EMBnet. journal. 2011 May 2;17(1):10.

60. Andrew S. FastQC: A Quality Control Tool for High Throughput Sequence Data. 2010.

61. Bolyen E, Rideout JR, Dillon MR, Bokulich NA, Abnet CC, Al-Ghalith GA, et al. Reproducible, interactive, scalable and extensible microbiome data science using QIIME 2. Nat Biotechnol. 2019 Aug 24;37(8):852–7.

62. Fettweis JM, Serrano MG, Sheth NU, Mayer CM, Glascock AL, Brooks JP, et al. Species-level classification of the vaginal microbiome. BMC Genomics. 2012 Dec 17;13(S8):S17.

63. Brooks M, Kristensen K, van Benthem K, Magnusson A, Berg C, Nielsen A, et al. Modeling zero-inflated count data with glmmTMB. bioRxiv. 2017;132753.

64. Hartig F. DHARMa: residual diagnostics for hierarchical (multi-level/mixed) regression models. 2020.

65. Lenth R, Buerkner P, Herve M, Love J, Riebl H, Singmann H. Estimated Marginal Means, aka Least-Squares Means. 2020.

66. Fernandes AD, Macklaim JM, Linn TG, Reid G, Gloor GB. ANOVA-Like Differential Expression (ALDEx) Analysis for Mixed Population RNA-Seq. Parkinson J, editor. PLoS One. 2013 Jul 2;8(7):e67019.

67. Fernandes AD, Reid JN, Macklaim JM, McMurrough TA, Edgell DR, Gloor GB. Unifying the analysis of high-throughput sequencing datasets: characterizing RNA-seq, 16S rRNA gene sequencing and selective growth experiments by compositional data analysis. Microbiome. 2014;2(2):15.

68. Gloor GB, Macklaim JM, Pawlowsky-Glahn V, Egozcue JJ. Microbiome datasets are compositional: And this is not optional. Front Microbiol. 2017;8(NOV):1–6.

69. Palarea-Albaladejo J, MartÍn-Fernández JA. zCompositions — R package for multivariate imputation of left-censored data under a compositional approach. Chemom Intell Lab Syst. 2015 Apr;143:85–96.

70. Oksanen J, Blanchet GF, Friendly M, Kindt R, Legendre P, McGlinn D, et al. vegan: Community Ecology Package. 2019.

71. Schwager E, Huttenhower C. banocc: Bayesian ANalysis Of Compositional Covariance. 2020.

72. Epskamp S, Cramer AOJ, Waldorp LJ, Schmittmann VD, Borsboom D. qgraph : Network Visualizations of Relationships in Psychometric Data. J Stat Softw. 2012;48(4).

