## Supplementary material for "The effect of secretor status and the vaginal microbiome on birth outcome"

### Supplementary Materials

### Supplementary Figures

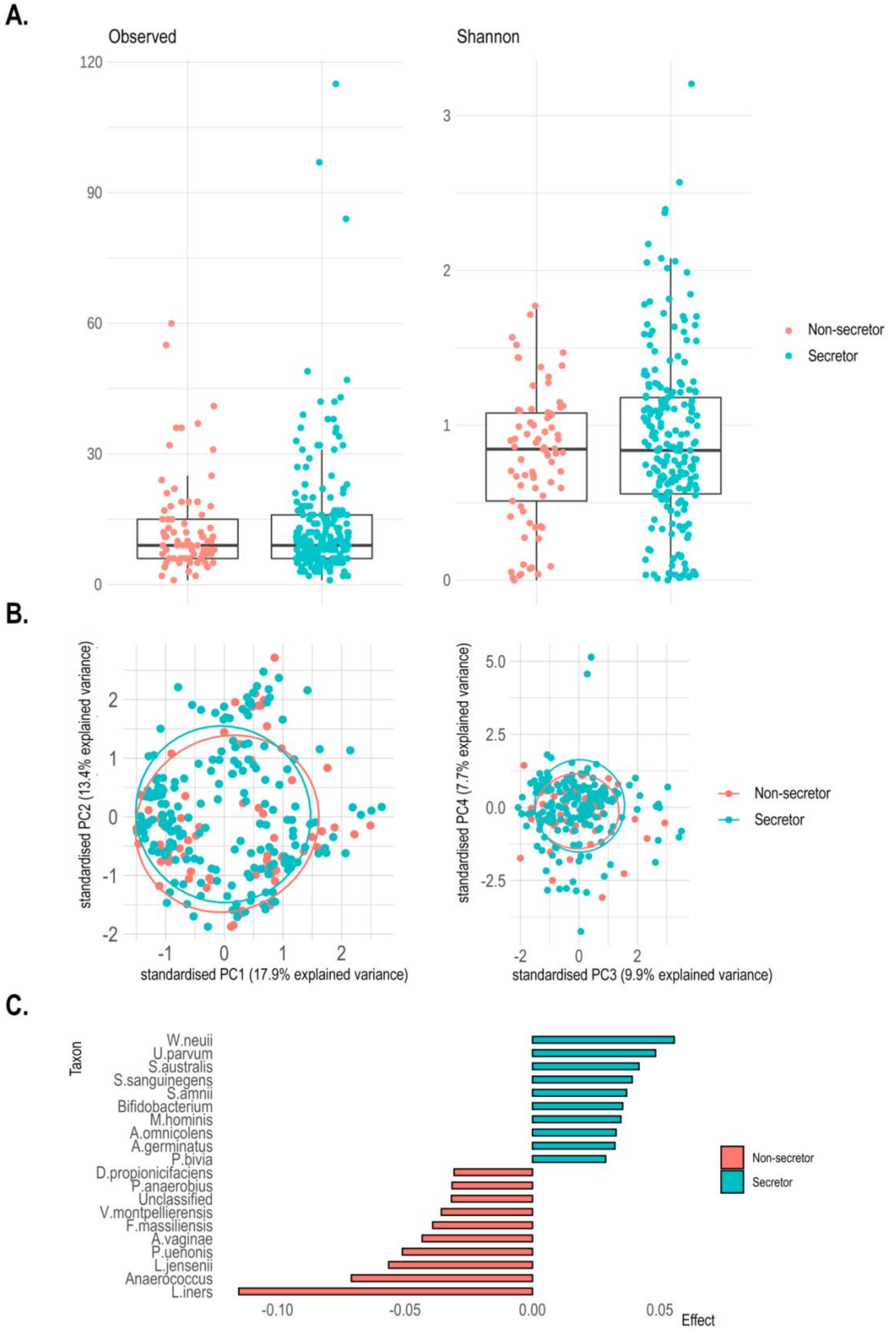

**Figure S1**. Secretors and non-secretors have similar vaginal microbiota profiles in early pregnancy. A. Distribution of diversity estimates of the vaginal microbiota in secretors and non-secretors in early pregnancy. B. PCA of the vaginal microbial community in early pregnancy shows no significant difference (first four principal components plotted). C. Differential Abundance Analyses indicated only small and non-significant differences between secretors and non-secretors in the top 10 microbial taxa by effect size, in the early pregnancy microbiota.

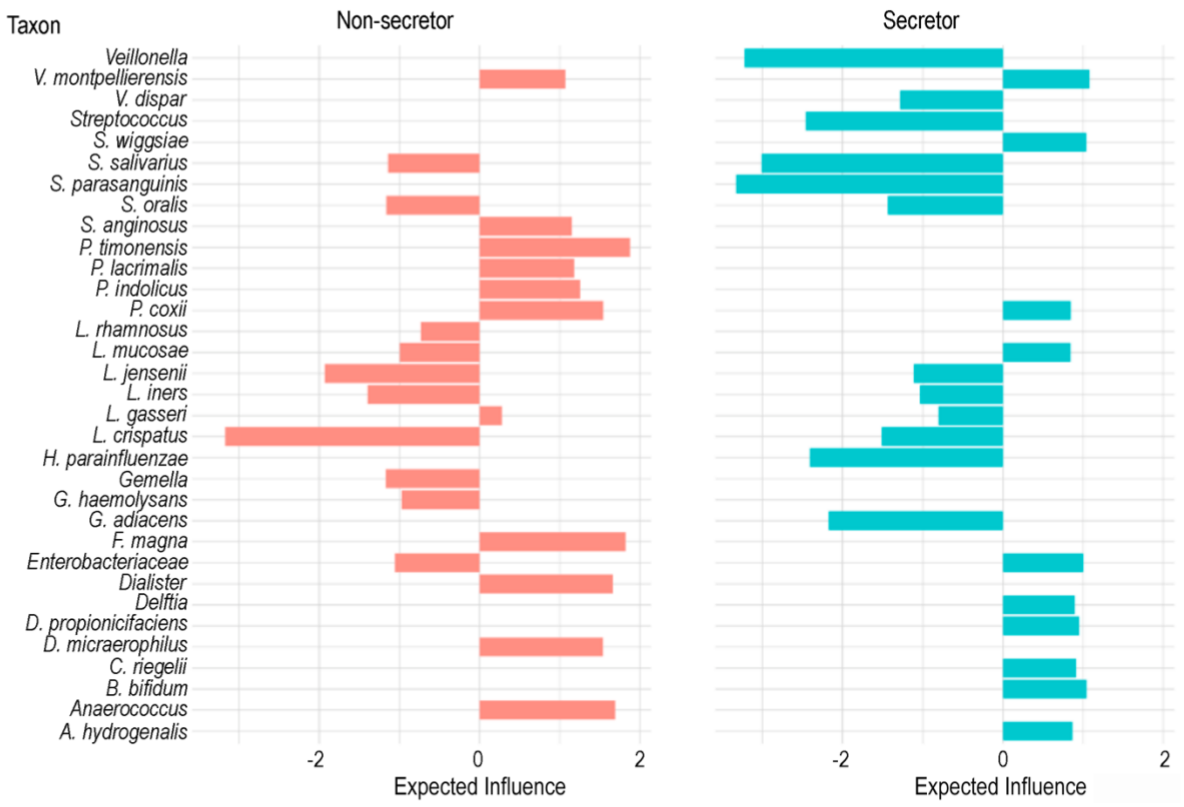


**Figure S2.** Distribution of nodes with the most positive and negative Expected Influence values (scaled) in non-secretors and secretors calculated for vaginal microbiome samples taken in early pregnancy.

### Supplementary Tables

**Table S1.** Clinical characteristics and inferred secretor status of the study cohort. Sample sizes and percentages (in brackets) are shown. Women (n=313) were sampled up to three times throughout pregnancy (Early: 63 – 130 days [n=287, median of 104.5], Mid:118 – 180 days [n=281, median of 150] and Late: 163 – 252 [n=253, median of 215]). Slightly irregularly timing of clinical visits led to a minor overlap between the mid and late sampling periods for a small number of patients. Secretor status was inferred from the presence of previously documented nonsense and missense mutations (heterozygotes were assumed to be secretors).*1 Preterm birth. *2 Mid trimester loss. *3 European ethnic group includes all individuals who identified themselves as ethnically white as well as those of mixed european ancestry.

| Variable | | Secretor | Non-secretor |
|---|---|---|---|
| Mean±SD gestational length in days | | 261.62 ±27.43 | 262.21 ±28.04 |
| Previous PTB[*1]/MTL[*2] | Yes | 114 (50.4) | 41 (47.1) |
| | No | 112 (49.6) | 46 (52.9) |
| Previous cervical excisional treatment | Yes | 75 (33.2) | 27 (31.0) |
| | No | 151 (66.8) | 60 (69.0) |
| Cervical stitch | Yes | 101 (44.7) | 36 (41.4) |
| | No | 125 (55.3) | 51 (58.6) |
| BMI | <18.5 | 4 (1.8) | 0 (0) |
| | 18.5-24.99 | 120 (53.1) | 50 (57.5) |
| | 25.0-29.99 | 67 (29.6) | 22 (25.3) |
| | >30.0 | 35 (15.5) | 15 (17.2) |
| Ethnicity | Afro-Caribbean | 34 (15.0) | 18 (20.7) |
| | Central-South Asia | 21 (9.3) | 5 (5.7) |
| | East Asia | 2 (0.9) | 3 (3.4) |
| | European[*3] | 129 (57.1) | 52 (59.8) |
| | Middle East/North Africa | 11 (4.9) | 2 (2.3) |
| | Mixed/Other | 29 (12.8) | 7 (8.0) |

**Table S2.** Mean gestational length for secretors and non-secretors and their vaginal microbiomes through pregnancy. Sample sizes are included in brackets.

| Microbiome | Early | | Mid | | Late | |
|---|---|---|---|---|---|---|
| | Secretor | Non-secretor | Secretor | Non-secretor | Secretor | Non-secretor |
| *Lactobacillus* dominated | 261.78 (163) | 265.90 (62) | 262.46 (166) | 263.80 (62) | 266.09 (141) | 267.59 (50) |
| *Lactobacillus* depleted | 264.34 (47) | 245.47 (15) | 265.58 (40) | 257.69 (13) | 261.73 (48) | 266.57 (14) |
| CST 1 | 263.95 (83) | 269.42 (31) | 264.35 (93) | 267.78 (36) | 266.79 (77) | 267.63 (27) |
| CST 2 | 264.70 (23) | 249.33 (6) | 264.63 (24) | 248.5 (6) | 258.5 (14) | 262 (5) |
| CST 3 | 257.89 (56) | 260.5 (30) | 258.06 (48) | 261.81 (17) | 265.61 (49) | 268.6 (21) |
| CST 4 | 264.06 (35) | 232.17 (6) | 266.86 (28) | 256.13 (8) | 264.49 (37) | 266.6 (5) |
| CST 5 | 262.62 (13) | 278 (4) | 261.23 (13) | 259.13 (8) | 259.83 (12) | 267.17 (6) |

**Table S3.** Analysis of deviance results from generalised linear mixed effects modelling (GLMM) of gestational length (days) using *Lactobacillus* status (at three timepoints in pregnancy) and ethnicity as a random effect. [†]significant in the model at $\alpha$=0.05. *1 Preterm birth. *2 Mid trimester loss.

| Model | Variable | Chisq | DF | P-value |
|---|---|---|---|---|
| Early | Age | 0.113 | 1 | 0.737 |
| | BMI | 3.431 | 3 | 0.330 |
| | Previous PTB[*1]/MTL[*2] | 13.185 | 1 | $2.821\times10^{-4\dagger}$ |
| | Cervical stitch | 14.283 | 1 | $1.573\times10^{-4\dagger}$ |
| | Previous cervical excisional treatment | 7.806 | 1 | $5.207\times10^{-3\dagger}$ |
| | Secretor × *Lactobacillus* | 5.117 | 1 | 0.024[†] |
| Mid | Age | 0.560 | 1 | 0.454 |
| | BMI | 0.211 | 3 | 0.976 |
| | Previous PTB[*1]/MTL[*2] | 10.314 | 1 | 0.001[†] |
| | Cervical stitch | 16.334 | 1 | $5.31\times10^{-5\dagger}$ |
| | Previous cervical excisional treatment | 7.576 | 1 | 0.006[†] |
| | Secretor × *Lactobacillus* | 0.711 | 1 | 0.399 |
| Late | Age | 1.322 | 1 | 0.250 |
| | BMI | 2.845 | 3 | 0.416 |
| | Previous PTB[*1]/MTL[*2] | 5.631 | 1 | 0.018[†] |
| | Cervical stitch | 6.090 | 1 | 0.014[†] |
| | Previous cervical excisional treatment | 6.550 | 1 | 0.010[†] |
| | Secretor × *Lactobacillus* | 0.474 | 1 | 0.491 |

**Table S4.** Analysis of deviance results from GLMMs of gestational length in days using Community State Type (CST) (with ethnicity as a random effect) highlighting covariates that are significant explanatory variables in the models. †Significant in the model at $\alpha$=0.05. *1 Preterm birth. *2 Mid trimester loss.

| Model | Variable | Chisq | DF | P-value |
|---|---|---|---|---|
| Early | Age | 0.477 | 1 | 0.490 |
| | BMI | 4.877 | 3 | 0.181 |
| | Previous PTB[*1]/MTL[*2] | 11.722 | 1 | $6.177\times10^{-4\dagger}$ |
| | Cervical stitch | 12.941 | 1 | $3.215\times10^{-4\dagger}$ |
| | Previous cervical excisional treatment | 8.963 | 1 | 0.003† |
| | Secretor × CST | 12.021 | 1 | 0.017† |
| Mid | Age | 1.791 | 1 | 0.181 |
| | BMI | 2.193 | 3 | 0.533 |
| | Previous PTB[*1]/MTL[*2] | 8.334 | 1 | 0.004† |
| | Cervical stitch | 10/279 | 1 | 0.001† |
| | Previous cervical excisional treatment | 7.565 | 1 | 0.006† |
| | Secretor × CST | 11.008 | 8 | 0.201 |
| Late | Age | 1.107 | 1 | 0.293 |
| | BMI | 4.228 | 3 | 0.238 |
| | Previous PTB[*1]/MTL[*2] | 4.529 | 1 | 0.033† |
| | Cervical stitch | 8.692 | 1 | 0.003† |
| | Previous cervical excisional treatment | 6.676 | 1 | 0.010† |
| | Secretor × CST | 4.488 | 8 | 0.811 |

**Table S5. Unstandardised coefficients (*b*), standard error (SE) and 95% confidence intervals (CI) from GLMMs of gestational length and incorporating ABO status (as an additional independent variable).** Blood group A, Secretors with *Lactobacillus* dominated microbiota are baseline in the model. $^{*1}$ Preterm birth. $^{*2}$ Mid-trimester loss. * p<0.05. ** p<0.01. *** p<0.001.

| | Regression model Early | | Mid | | Late | |
|---|---|---|---|---|---|---|
| | b (SE) | CI | b (SE) | CI | b (SE) | CI |
| Intercept | 3.09 (0.29) | 2.52,3.67 | 2.99 (0.30) | 2.41,3.60 | 2.95 (0.31) | 2.35,3.55 |
| Age | 0.002 (0.01) | -0.01,0.02 | 0.005 (0.01) | -0.01,0.02 | 0.01 (0.01) | -0.01,0.02 |
| BMI (<18.5) | -0.10 (0.41) | -0.92,0.68 | 0.06 (0.34) | -0.60,0.67 | 0.05 (0.38) | -0.68,0.79 |
| BMI (25.0-29.99) | 0.07 (0.09) | -0.11,0.25 | -0.04 (0.1) | -0.22,0.15 | 0.07 (0.1) | -0.13,0.26 |
| BMI (>30.0) | -0.18 (0.13) | -0.43,0.07 | -0.11 (0.12) | -0.34,0.12 | -0.12 (0.12) | -0.36,0.13 |
| Cervical stitch | 0.30 (0.09)$^{***}$ | 0.13,0.48 | 0.31 (0.08)$^{***}$ | 0.14,0.47 | 0.20 (0.09)$^{*}$ | 0.03,0.36 |
| Previous PTB$^{*1}$/MTL$^{*2}$ | 0.41 (0.1)$^{***}$ | 0.21,0.61 | 0.37 (0.1)$^{***}$ | 0.18,0.57 | 0.25 (0.1)$^{*}$ | 0.05,0.45 |
| Previous cervical excisional treatment | -0.23 (0.1)$^{*}$ | -0.43,-0.02 | -0.20 (0.1)$^{*}$ | -0.41,-0.02 | -0.23 (0.1)$^{*}$ | -0.43,-0.03 |
| ABO (AB) | -0.13 (0.21) | -0.55,0.29 | -0.06 (0.2) | -0.46,0.34 | -0.03 (0.21) | -0.44,0.38 |
| ABO (B) | 0.22 (0.12) | -0.02,0.46 | 0.31 (0.12)$^{**}$ | 0.08,0.55 | 0.13 (0.12) | -0.10,0.37 |
| ABO (O) | 0.04 (0.1) | -0.15,0.23 | -0.04 (0.1) | -0.22,0.15 | -0.06 (0.1) | -0.25,0.13 |
| Secretor × *Lactobacillus* | 0.54 (0.23)$^{*}$ | 0.08,1.0 | 0.27 (0.24) | -0.27,0.67 | -0.11 (0.23) | -0.57,0.34 |
